# Lung cancer pathway inequalities for adults with severe mental health conditions: A mixed-methods analysis of barriers to screening and care pathways in South East London

**DOI:** 10.64898/2026.06.08.26355143

**Authors:** Gracie Tredget, Maria Milenova, Reanne Parkash, Raymond McGrath, Mark Edwards, Siobhan Gee, Wesley Pigg, Darek Karwacki, Charlie Costa, Sanna Shafique, Mark Adams, Jackie Waghorn, David I’Anson, Amy Ronaldson, Kate Haire, Charmi Githuku, Ed Beveridge, Julie Williams

**Affiliations:** South London and Maudsley NHS Foundation Trust, London, SE5 8AZ, UK; Centre for Global Mental Health, Institute of Psychiatry, Psychology and Neuroscience, King’s College London, 16 De Crespigny Park, London, SE5 8AF, UK Health Services and Population Research, King’s College London; Department of Addictions, Institute of Psychiatry, Psychology and Neuroscience, King’s College London, 4 Windsor Walk, SE5 8BB; Department of Basic and Clinical Neuroscience, Institute of Psychiatry, Psychology and Neuroscience, King’s College London; Woolwich Service Users Project, 107 Brookhill RD, SE18 6BJ; Mind & Body Programme, King’s Health Partners, SE1 9RT; Clinical and Care Professional Lead for Cancer, Lambeth, NHS South East London Integrated Care Board, 160 Tooley Street, London, SE1 2TZ, UK; Guy’s & St. Thomas’ NHS Foundation Trust, Westminster Bridge Road, London, SE1 7EH, UK; NHS South East London Integrated Care Board, 160 Tooley Street, London, SE1 2TZ, UK; Centre for Mental Health Policy and Evaluation, Health Services and Population Research Department, IoPPN, King’s College London, SE5 8AF; South East London Cancer Alliance, London SE1 9RT, UK; Central and North West London NHS Foundation Trust, London NW1 3AX; School of Communities and Education, Northumbria University, Newcastle upon Tyne, UK

**Keywords:** severe mental health conditions, lung cancer, health inequalities, cancer screening, diagnostic pathway, mixed methods

## Abstract

**Background:** Adults with severe mental health conditions (often referred to as severe mental illness – SMI) experience 15–20-year mortality gap relative to the general population, with lung cancer a significant contributor. National cancer policy targets earlier diagnosis but does not explicitly address how pathways function for this group.

**Aims:** This study aimed to describe lung cancer risk, prevalence, screening eligibility, referral activity and diagnostic pathway performance for adults with SMI in South East London (SEL), and to examine where along the pathway inequalities arise.

**Methods:** Co-designed with experts with lived experience and voluntary sector, this exploratory mixed-methods service evaluation combined quantitative analysis of routinely collected data from the Quality Outcomes Framework (QOF), SMI Register and Cancer Waiting Times Record (April 2023-March 2024) with semi-structured qualitative interviews (n=11 clinical staff) and focus groups (n=6 adults with lived experience of SMI). Quantitative and qualitative data were analysed using descriptive statistics and framework-based thematic analysis respectively, and findings were integrated using a joint display approach, organised by the Consolidated Framework for Implementation Research (CFIR).

**Results:** Lung cancer prevalence was approximately double among adults with SMI (0.17% vs 0.09% in the general population). Despite Urgent Suspected Cancer (USC) referral rates being more than twice as high in the SMI population (63 vs 28 per 100,000), fewer cancers were detected via planned general practice (GP) routes (11% vs 20%), the 28-day Faster Diagnosis Standard was not met for any SMI patient diagnosed with lung cancer during the study period; overall FDS performance was 76% in the SMI population compared with 84% in the general population; and appointment non-attendance was more than double that in the general population (6% vs 3%). Qualitative findings identified individual, service and system-level mechanisms – including stigma, diagnostic overshadowing, fragmented coordination, and rigid pathway protocols – that compound disadvantage across lung cancer pathway stages.

**Conclusions:** Inequality in lung cancer outcomes for adults with SMI accumulates across the pathway rather than arising at a single point of failure. Addressing this requires proportionate adaptations within existing cancer pathways, alongside routine reporting of cancer outcomes stratified by SMI population.

**Significance of this study:** **What is already known on this subject?**

- People with severe mental health conditions (often referred to as severe mental illness - SMI) experience substantially worse cancer outcomes, including higher mortality from lung cancer.
- Inequalities are driven by multi-level barriers, including reduced access to screening, diagnostic delay, and fragmented care across physical and mental health services.
- However, there is limited empirically grounded evidence on how these barriers operate within lung cancer pathways, and how they can be addressed in practice.

**What are the new findings?**

- This study identifies multi-level barriers and enablers to the National Health Service Lung Cancer Screening (NHS LCS) programme and care for people with SMI, spanning individual, service, and system domains.
- Barriers include fragmentation between services, unclear care coordination, and challenges navigating screening pathways, even when eligibility is met.
- Findings are theoretically informed by three implementation science frameworks and validated through multi-stakeholder engagement, including clinicians and people with lived experience.
- The study generates co-produced, actionable recommendations to address these barriers across the pathway.

**How might this impact on clinical practice in the foreseeable future?**

- Provides an evidence-informed basis for redesigning lung cancer pathways to improve access, engagement, and continuity of care for people with SMI.
- Supports targeted service and system interventions to address identified barriers and improve early diagnosis.
- Offers practical, co-produced recommendations for clinicians and system leaders working to reduce inequalities.
- Findings have direct relevance to ongoing national and regional efforts to reduce cancer inequalities and improve outcomes for people with SMI.

## Introduction

Lung cancer remains the leading cause of cancer-related death in the United Kingdom (UK) and a major focus of national cancer policy (Cancer Research UK, 2025). In 2019, the National Health Service (NHS) Long Term Plan set out the ambition of diagnosing 75% of cancers at Stage I-II (NHS England, 2025a). The National Cancer Plan for England (2026-2035) includes lung cancer specific initiatives alongside recognition that wider physical, mental health and social needs must be addressed for patients within cancer pathways (Department of Health & Social Care, 2026). National programmes including the NHS Lung Cancer Screening (LCS) programme, Non-site specific symptom (NSS) services, and Cancer Waiting Time (CWT) standards have contributed to meaningful improvements in earlier diagnosis in the general population (Kerrison et al., 2023). However, these approaches are largely designed around age-based eligibility, symptom presentation, and standardised referral pathways, and do not explicitly account for the needs of adults living with severe mental health conditions (often referred to as severe mental illness and throughout this paper, SMI) (Tuschick et al., 2024).

Adults with SMI (defined here as individuals with schizophrenia, schizoaffective disorder, bipolar disorder, or other causes of psychosis) experience significantly poorer cancer outcomes than the general population. A nationwide cohort study in the UK found a 64% higher all-cause mortality rate following cancer diagnosis for adults with SMI compared with those without (Launders et al., 2022), attributed to delayed diagnosis, more advanced cancer stage at diagnosis and reduced engagement with cancer care services. A recent systematic review also found consistent evidence that cancer is more frequently diagnosed at an advanced stage in adults with psychotic disorders, suggesting that more effective support for accessing screening and Urgent Suspected Cancer (USC) referral pathways could improve outcomes (Wootten et al., 2022).

Smoking is the single largest modifiable contributor to the mortality gap experienced by adults with SMI (Campion et al., 2023). National estimates indicate smoking prevalence among people with SMI is between 40% and 90%, depending on the setting, whereby adults with SMI typically smoke more heavily and for longer than the general population (Brose et al., 2020). Lung cancer, the third most common cancer in the UK, is directly linked to smoking in 72% of cases. Incidence rates of lung cancer are 174% higher in populations experiencing social deprivation (Cancer Research UK, 2025). Adults with SMI are also disproportionately concentrated in deprived communities where smoking prevalence is high. For example, in South East London (SEL) nearly two-thirds of people on SMI registers live in the most deprived 40% of neighbourhoods (Tredget and Williams, 2023).

In 2021, the Core20PLUS5 framework identified adults with SMI as a priority population for reducing health inequalities, including in cancer (NHS England, 2021). Despite this growing policy attention, there is limited empirical evidence describing how cancer pathways function in practice for this population. A recent national expert roundtable examined barriers to early lung cancer diagnosis in people with mental health conditions, identifying systemic barriers including diagnostic overshadowing, fragmented services, passive screening invitation models, and the absence of SMI-specific pathways (Bristol Myers Squibb, 2026). The roundtable, while valuable, was based on expert consensus rather than primary data, used a broad definition of mental health conditions, and was not co-designed with or included evidence from people with lived experience of SMI. How lung cancer pathways function in practice for adults with SMI (e.g., where along the pathway inequalities emerge, and how the pathway is navigated by both staff and people with lived experience) remains largely unanswered (Millman et al., 2016, Tuschick et al., 2024). This study was undertaken to address these gaps and focus on areas for improvement across lung cancer pathways that could support better access, adherence and outcomes for adults with SMI.

### Aims

The aims of this study were to: (1) describe lung cancer risk, prevalence (i.e., derived from long-term incidence and survival rates to provide the total cancer burden for this population), screening uptake, referral activity, and diagnostic pathway performance for adults with SMI in SEL; and to (2) identify the barriers that prevent access and adherence to lung cancer symptomatic diagnostic pathways and at what point in the pathway inequalities arise. The study aimed to provide actionable, system-level evidence to inform actionable and equitable lung cancer pathway improvement for the SMI population.

## Methods

### Study design and setting

This exploratory mixed-methods service evaluation was conducted across the six boroughs of SEL (Bexley, Bromley, Greenwich, Lambeth, Lewisham, and Southwark) and focused on the general practice (GP) registered SMI population within the region (approximately 1% of the general population in SEL, or 1 in 80 residents) as recorded in Quality Outcomes Framework (QOF), April 2023-March 2024. The study followed a sequential exploratory design (Creswell and Plano Clark, 2023) in which quantitative findings informed the qualitative investigation. Routinely collected quantitative data were integrated with qualitative data from interviews, focus groups, and co-produced workshops. This study received ethical approval from Health Research Authority (HRA) (number 25/LO/0523).

### Stakeholder Involvement

A bi-monthly multidisciplinary steering group was convened for the duration of the study, comprising system partners from primary care, mental health services, acute Trusts, voluntary, community and social enterprise (VCSE) organisations, public health providers, people with lived experience, and the South East London Cancer Alliance (SELCA). Its role was to support ongoing identification of barriers and inequalities, sharing best practice examples, identify scalable opportunities for change, and guide interpretation of emerging findings.

Lived experience was central to the study design. The study team partnered with Woolwich Service Users Project (WSUP), a local VCSE organisation, to support meaningful engagement of adults with lived experience of SMI who had accessed local lung cancer pathways. Two experts by experience were integral members of the project team and contributed to steering group meetings and to the co-design and delivery of the people with lived experience focus groups. Focus group materials, recruitment approaches, and reimbursement methods were co-produced with WSUP and the experts to ensure fair compensation, accessibility and inclusivity. Emerging findings were iteratively refined through two workshops involving people with lived experience and staff (Supplementary file 1), some of whom also contributed to co-writing the study outputs.

### Recruitment, data collection and analyses

#### Phase 1: Quantitative data – gathering routinely collected data from QOF SMI and Cancer Waiting Time registers

Anonymised and routinely collected data from primary care records was used for this study. The data derived from QOF SMI registers within general practice records and Cancer Waiting Times (CWT) collections. Data were linked, where possible, to local cancer activity and pathway information to examine population characteristics (e.g., age, gender, ethnicity, borough and deprivation decile), smoking status, lung cancer prevalence, referral and detection rates, diagnostic pathway indicators including the 28-day Faster Diagnosis Standard (FDS) (NHS England, 2025b), appointment non-attendance rates (Did Not Attend – DNAs), and NHS LCS eligibility. The population of adults with SMI was identified using QOF registers held in primary care, which capture individuals with schizophrenia, schizoaffective disorder, bipolar disorder, or other causes of psychosis, registered with a GP. All prevalence and rate estimates used the QOF-registered SMI population as the denominator. Data covered the period 1st April 2023-31st March 24 and were extracted on 31st March 2024. The dashboards used, in accordance with information governance requirements for the use of this data, apply “small number suppression”, whereby any values <5 were suppressed to minimise the risk of patient identification. These dashboards have been used with the explicit permission and rigorous oversight of the Report Sponsor within the SEL ICB Planned Care and Cancer Team, to ensure safe and appropriate use in line with local use policies.

#### Phase 2: Qualitative data – semi-structured interviews with staff and focus groups with people with lived experience of SMI

##### Recruitment and setting

Eleven clinical staff from across the lung cancer pathway in SEL were recruited, including staff from primary care, mental health services and emergency service departments (ED). Participants were identified using purposive sampling through the project team’s professional networks. Recruitment involved advertisement via email distributions and attendance at relevant forums across SEL to promote the study. Interested clinicians received a participant information sheet and consent form. Following written consent, semi-structured interviews were conducted virtually using MS Teams.

Six adults with lived experience of SMI who had accessed the lung cancer pathway within SEL were recruited to take part in in-person focus groups. Recruitment was led by WSUP, using a purposive sampling approach. Recruitment involved advertising through WSUP’s established networks, including its VSCE networks and direct communication with its members, using recruitment materials co-developed with the project’s monthly planning group. Additional outreach was undertaken via the project teams clinical networks and a local radio advertisement featuring the Chair of WSUP and experts by experience from the project team.

##### Analysis

Data were recorded and transcribed by MS Teams. Transcripts were reviewed, anonymised and coded by members of the research team (MM, RP, SS) in Excel. Analysis was primarily deductive and interpretive, guided by a hybrid framework integrating the Health Equity Implementation Framework (HEIF) (Woodward et al., 2021), the Consolidated Framework for Implementation Research (CFIR 2.0) (Damschroder et al., 2022), and selected domains from the Theoretical Domains Framework (TDF) (Cane et al., 2012). CFIR provided the preliminary structure for examining multi-level implementation determinants; HEIF was incorporated to ensure explicit attention to equity-relevant contextual and recipient factors that CFIR addresses less directly; and selected TDF domains enabling finer-grained examination of behavioural determinants at the individual level. This structure supported analysis across individual, service, and system levels while retaining sensitivity to equity determinants at each level.

Each transcript was coded by one researcher and reviewed by a second; discrepancies in interpretation were resolved through team review and discussion. Findings were organised within the integrated coding framework (Supplementary file 2) and iteratively refined through wider team discussions and through two stakeholder workshops, one with people with lived experience and the other with staff (see Stakeholder Involvement section). This iterative refinement ensured interpretations remained grounded in participant accounts while supporting cross-cutting synthesis across individual, service and system levels.

### Data Integration

Qualitative and quantitative findings were integrated using a joint display approach (Fetters et al., 2013), organised by CFIR 2.0 domains. Quantitative indicators of pathway performance (e.g., referral rates, FDS compliance) were mapped alongside the qualitative themes that spoke to the same domain, enabling the qualitative data to help explain the patterns observed in the quantitative findings. Points of convergence, divergence, and expansion between the two datasets were identified through team discussion. Consistent with sequential exploratory mixed methods approaches, integration focused on identifying convergent themes. Resulting findings are interpretive and exploratory, reflecting emergent insights rather than causal claims.

## Results

### Population characteristics

The QOF SMI registers in SEL contained 26,365 individuals coded as having SMI, representing 1% of the registered population. The largest SMI population cohorts by age were age groups 20-39 and 40-54 (Supplementary file 3). Nearly two-thirds (64%) of adults with SMI lived in the most deprived 40% of neighbourhoods in SEL.

### Smoking prevalence

Smoking prevalence in SEL for the SMI population was reported as 27%, compared with 4% reported in the non-SMI population in primary care at the time of reporting, representing an approximately six-fold difference. Smoking prevalence by age band and SMI status is presented in Table 1. Prevalence was highest among working age adults with SMI, peaking at 26% in those aged 40-54 (versus 5% in non-SMI peers) and 34% in those aged 55-74 (versus 8%). Approximately one in four adults with SMI in SEL were recorded as current smokers, compared with approximately one in twenty-five in the non-SMI population.

**Table 1.**
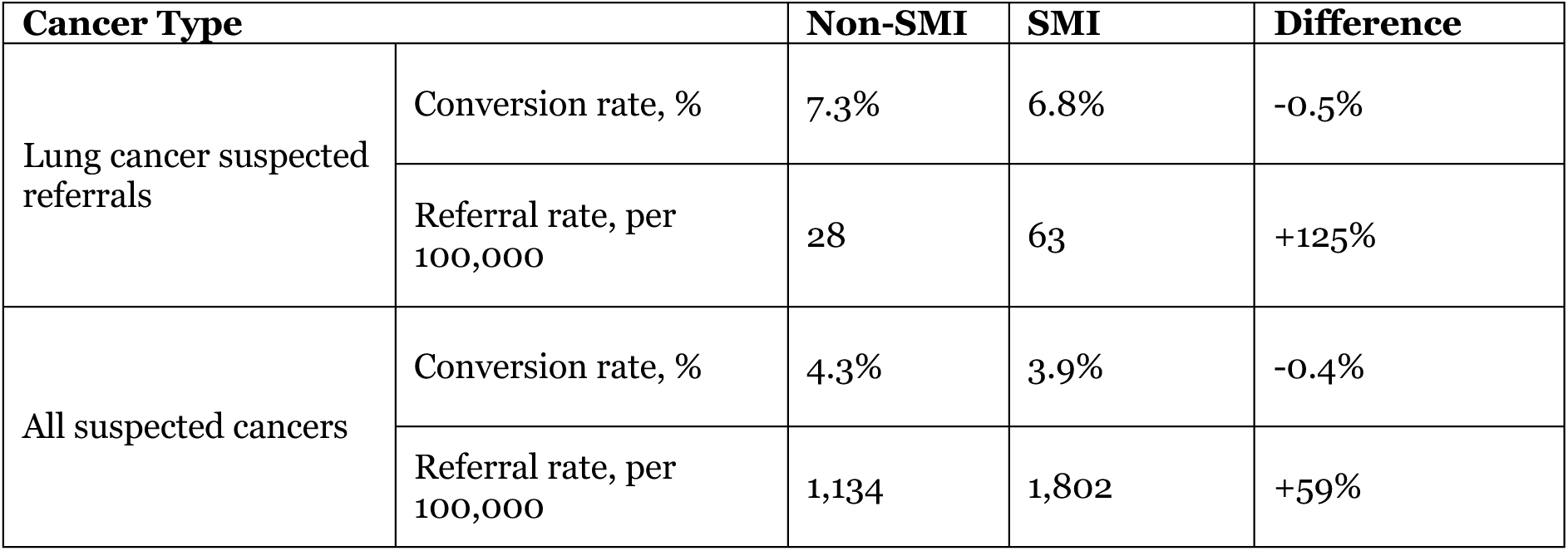
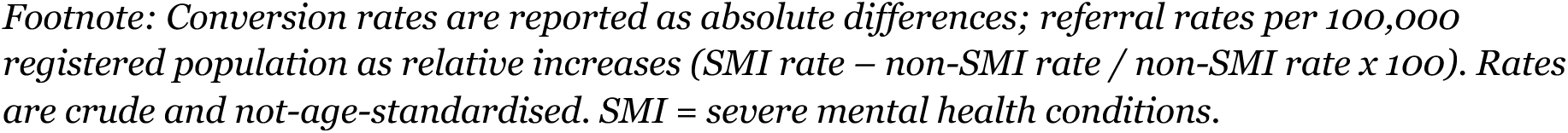
Conversion to diagnosis rate of Urgent Suspected Cancer (USC) referrals 2023/24 and referral rates per 100,000 for all-cancers, by SMI status.

| Cancer Type |  | Non-SMI | SMI | Difference |
| --- | --- | --- | --- | --- |
| Lung cancer suspected referrals | Conversion rate, % | 7.3% | 6.8% | -0.5% |
|  | Referral rate, per 100,000 | 28 | 63 | +125% |
| All suspected cancers | Conversion rate, % | 4.3% | 3.9% | -0.4% |
|  | Referral rate, per 100,000 | 1,134 | 1,802 | +59% |

Footnote: Conversion rates are reported as absolute differences; referral rates per 100,000 registered population as relative increases ( $\text{SMI rate} - \text{non-SMI rate} / \text{non-SMI rate} \times 100$ ). Rates are crude and not-age-standardised. SMI = severe mental health conditions.

### Lung cancer and all-cancer prevalence

Cancer incidence data stratified by SMI diagnosis were not available, therefore prevalence (point-in-time count of people living with a diagnosis) is reported. Because prevalence reflects both incidence and survival, and survival is known to be poorer in people with SMI, true incidence in this population is likely to be greater than the prevalence figures suggest. Lung cancer prevalence was 0.17% in the SMI population compared with 0.09% in the non-SMI population, indicating that lung cancer was almost twice as prevalent (see Fig. 2). All cancer prevalence was higher in the SMI population (4% versus 3%), with lung cancer accounting for a greater proportional share of the total cancer burden among adults with SMI compared to the general population, representing a 31% higher proportional contribution.

**Figure 1.**
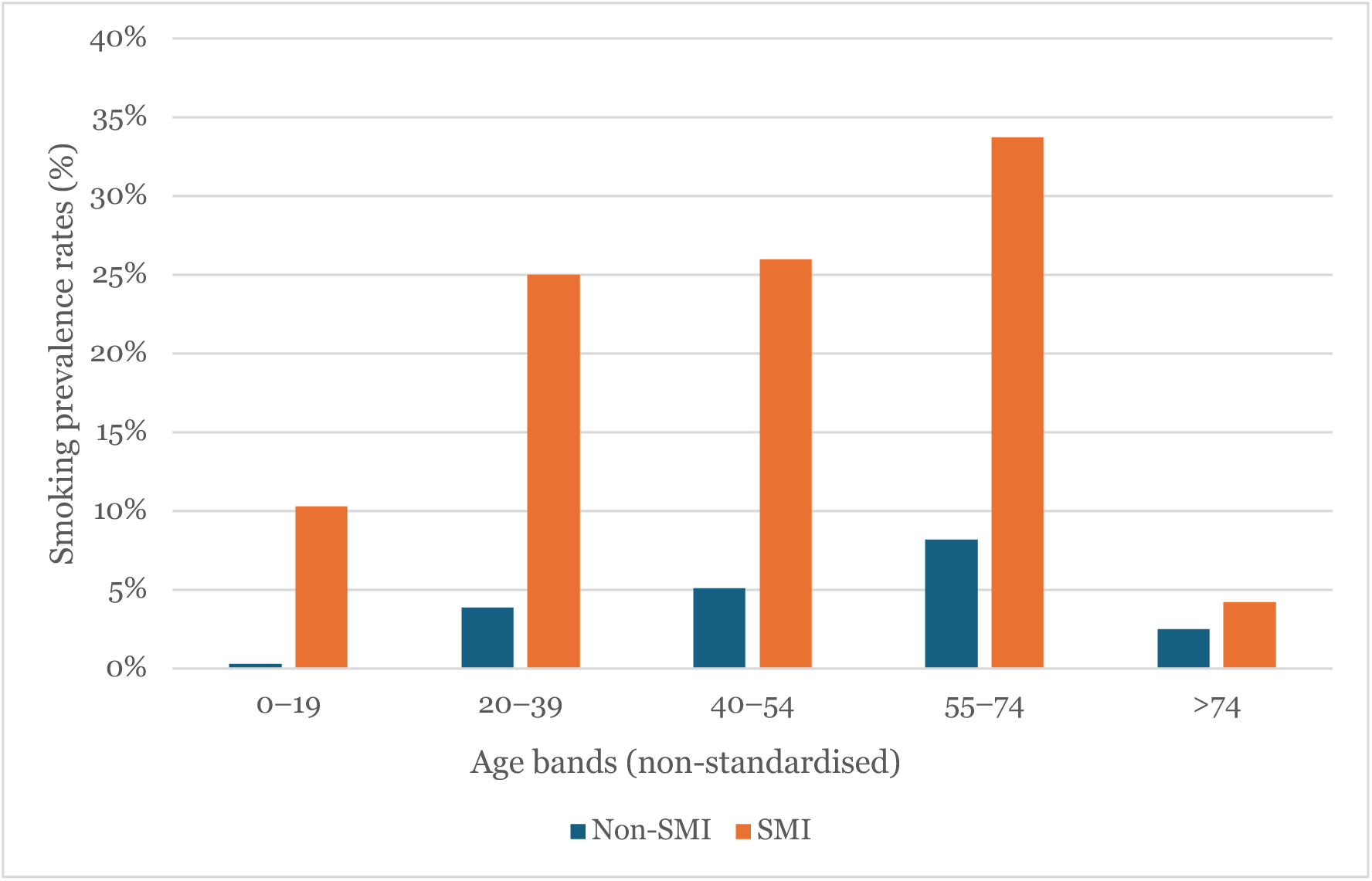
Smoking prevalence by age and SMI status. Smoking prevalence by age and severe mental health condition as recorded in South East London General Practice registers, snapshot 31 March 2024. Non-standardised age bands were used with a logic applied to see lung screening age bands within pre-existing age bands on the available registers. Although smoking among <18s is recorded, the meaningful comparison starts from adulthood. SMI = severe mental health condition.

**Figure 2.**
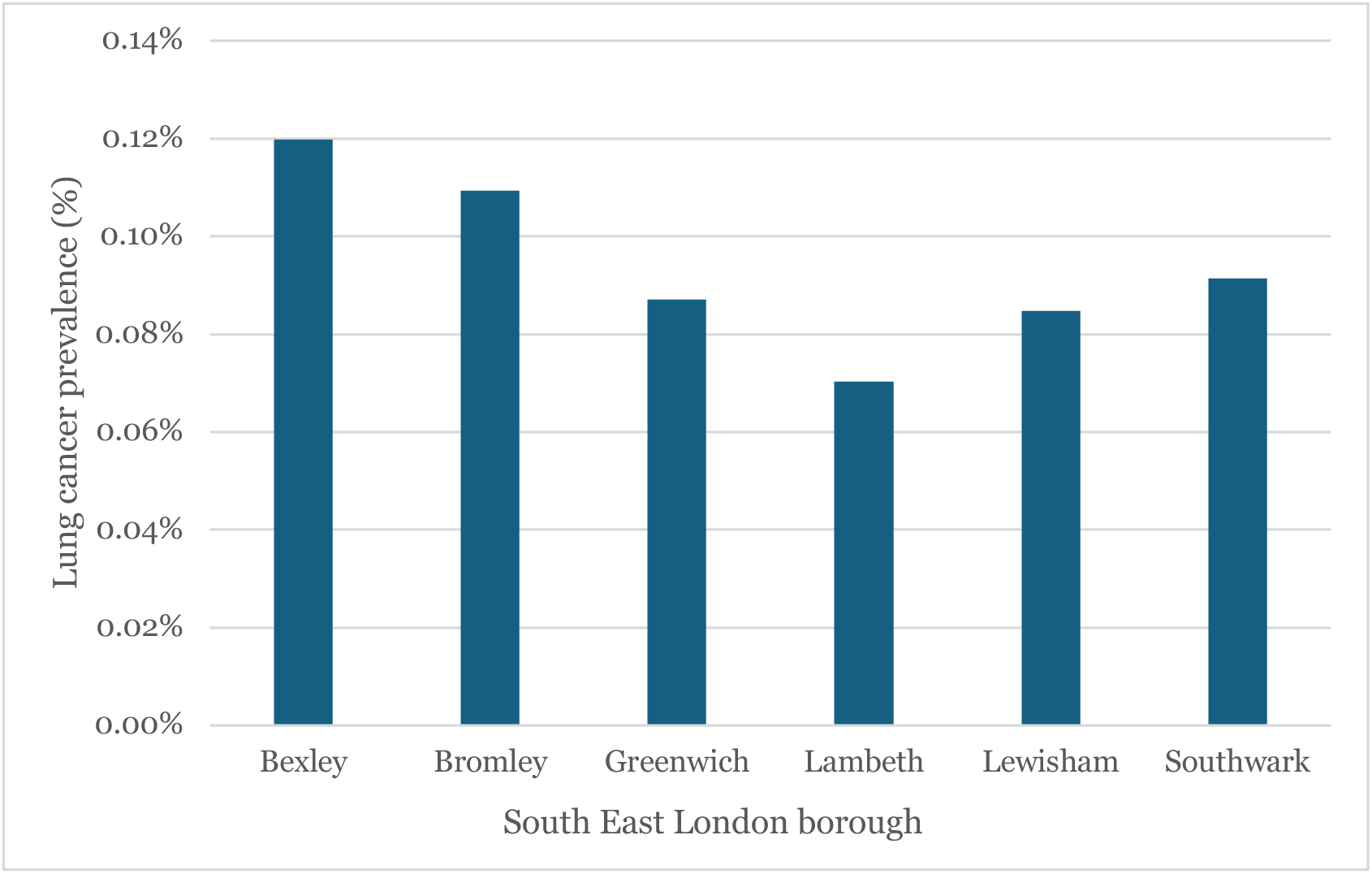
Borough variation in lung cancer prevalence by SMI status. Lung cancer prevalence by borough and SMI status, as recorded in South East London General Practice (SEL GP) registers, snapshot 31 March 2024. Prevalence >1 indicates higher prevalence in the SMI population. SMI = severe mental health conditions.

However, in either case, it was not possible to determine the prevalence of cancer stage as this data was not available at the time of reporting, and therefore it was not possible to determine how cancer stage may vary in prevalence rates between SMI and non-SMI populations.

### Detection route and referral rates

Eleven percent of lung cancer in the SMI population were diagnosed via GP referral for USC, compared with 20% in the non-SMI population. The available data did not include formal Routes to Diagnosis coding and could not distinguish between diagnosis via emergency presentation, screening, incidental findings, and other pathways. Despite the lower detection rate via the USC pathway for the SMI population, suspected cancer referral rates were substantially higher in the SMI population overall: Lung cancer referral rates were more than double compared to the general population (63 versus 28 per 100,000). However, referral to diagnosis conversion rates were slightly lower in the SMI population for both lung cancer and all cancers (by 0.5 and 0.4 percentage points respectively), indicating that higher referral volumes did not translate into proportionally more lung cancer diagnoses for the SMI population despite higher referral rates than the general population (see Table 1).

### Diagnostic timelines

None of the patients with SMI referred on an USC pathway who received a lung cancer diagnosis met the 28-day FDS (FDS is a national standard requiring that patients with urgent cancer referrals receive a diagnosis or have cancer ruled out within 28 days). This compared with 21% in the non-SMI population.

### Appointment non-attendance

Adults with SMI were almost twice as likely as those without SMI to not attend for outpatient lung cancer pathway appointments (DNA rate in adults with SMI: 6%, vs, 3% in the non-SMI population). Although absolute numbers were small (10 DNAs from 177 appointments), the pattern was consistent across all SEL boroughs.

### NHS LCS programme eligibility

Stratifying the data by age and SMI status (Figure 3) showed that the largest cohort of people with SMI are aged between 40 and 54. The LCS programme currently screens people aged 55-74, indicating a significant proportion of people with SMI are not eligible for LCS based on current screening thresholds.

**Figure 3.**
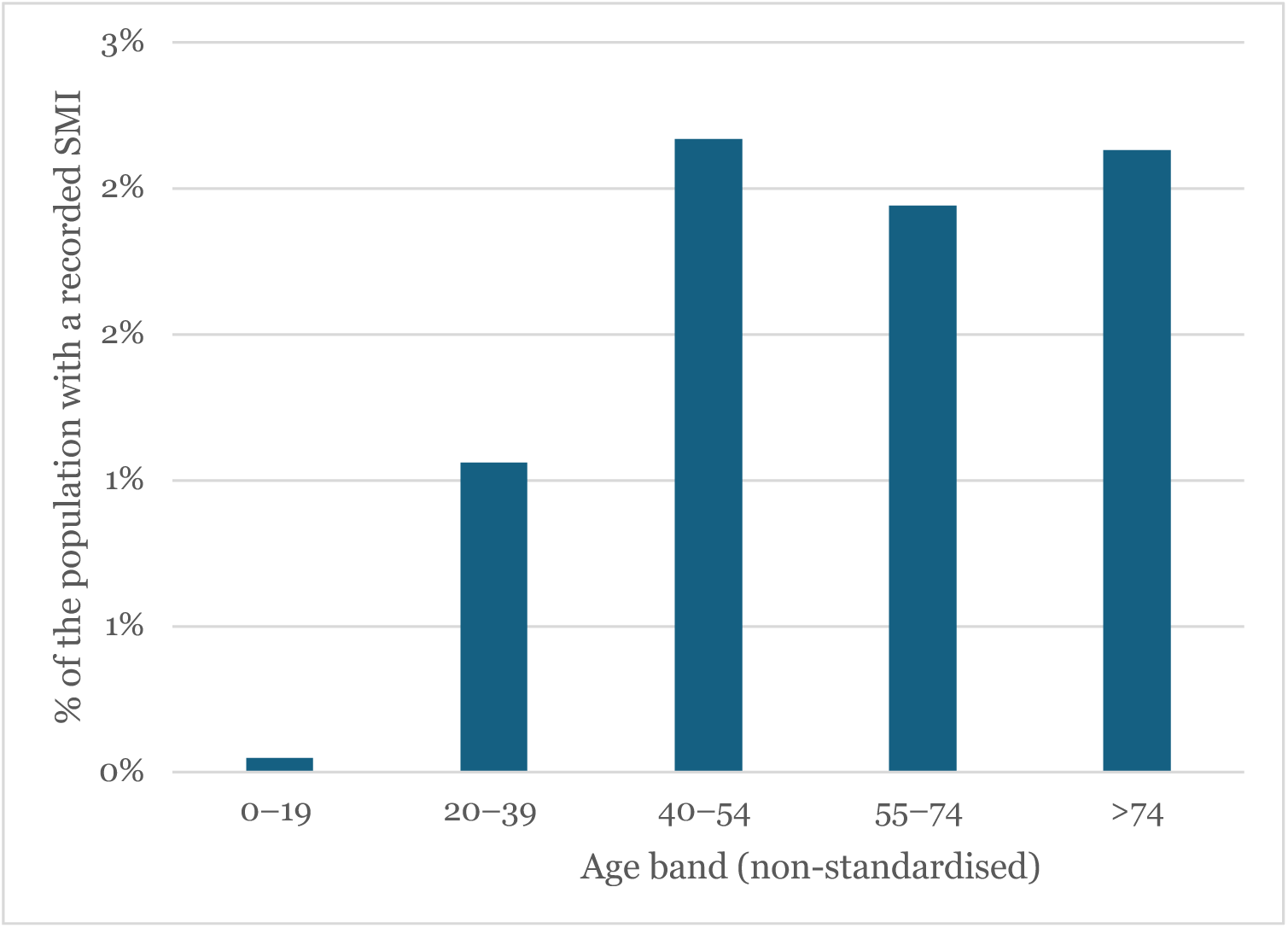
Age by SMI status. Age and severe mental health condition as recorded in South East London General Practice registers, snapshot 31 March 2024. Non-standardised age bands were used with a logic applied to see lung screening age bands within pre-existing age bands on the available registers. SMI – severe mental health conditions.

Among those with SMI who had a recorded lung cancer diagnosis, 67% were in the screening eligible age group, compared with 44% in the non-SMI population, reflecting the strong age-dependence of lung cancer development. Within the eligible age group, lung cancer prevalence among adults with SMI was 0.46% (approximately double the non-SMI rate of 0.23%). The implications for screening policy are explored further in the Discussion.

### Barriers and enablers across the pathway

Qualitative analysis of interviews and focus groups identified a set of interrelated barriers and enablers affecting access and adherence to NHS LCS pathways for adults living with SMI in SEL. These clustered across individual, service, and systems levels (see Table 2), with equity-related factors cutting across all stages of the pathway (Table 4). Supplementary file 4 describes focus groups and interview participant characteristics.

**Table 2:** Barriers across the lung cancer screening and diagnostic pathway for adults with SMI.

| Qualitative theme | Summary |
| --- | --- |
| **Individual-level barriers** |  |
| **Stigma and mistrust of healthcare services** | Stigma and mistrust delayed help-seeking and reduced willingness to disclose symptoms. Patients described feeling judged, particularly in relation to smoking, which led them to minimise or conceal symptoms. In one case, a patient on a forensic ward did not disclose coughing up blood for weeks before casually mentioning it in conversation. Mistrust was reinforced by prior experiences of being dismissed, patronised or criticised by healthcare professionals, and extended to medication and services perceived by some as punitive or controlling. Self-stigma and masculine identity (“bravado”) were identified as barriers to disclosure, particularly among male patients on forensic wards. |
| **Fear of diagnosis and avoidance behaviours** | Fear of diagnosis and avoidance behaviours led some patients to actively decline investigations. Staff described individuals refusing scans because psychotic symptoms instructed them not to engage, or because they felt that if treatment was not something they could face without support, there was no purpose in knowing. Coercive or fear-based messaging (for example, graphic images used to prompt engagement) was experienced as counterproductive and reinforced avoidance rather than encouraging uptake. |
| **Low health literacy and under-recognition of symptoms** | Low health literacy and under-recognition of symptoms contributed to delayed presentation. Persistent cough was frequently normalised among smokers, and patients had limited understanding of what investigations involved. Staff reported very low functional literacy among some patient groups, with a minority unable to read written health information, limiting engagement with standard educational materials and appointment correspondence. This was compounded by cognitive effects of SMI that affected memory and information retention. |
| **Day-to-day survival and competing life crises** | Emotional overwhelm, hopelessness, housing instability, food insecurity and poverty emerged as powerful motivational barriers. For many adults with SMI, managing day-to-day survival left limited capacity for engaging with preventative health. Staff observed that patients often presented to services for social rather than medical reasons, and that competing crises could make cancer screening feel remote and abstract. Expressions of hopelessness (“nothing to live for”) were reported by staff as directly undermining motivation to engage. |
| **Service-level barriers** |  |
| **Rigid non-attendance (DNA) and re-referral policies** | NHS policies typically close referrals after a set number of missed appointments, requiring patients to be re-referred through primary care to re-enter the pathway. Staff described individuals with SMI being disproportionately affected, illustrated by accounts of consultants personally contacting teams to keep a referral open for a patient struggling to attend early-morning scans. Fluctuating mental health, transport difficulties and confusion about appointment logistics can make consistent attendance genuinely difficult to sustain without active support, and rigid application of DNA policies compounds pathway disruption for those least able to navigate re-entry. |
| **Poorly adapted communication, correspondence and digital access** | Standard pathway communication relies on written letters containing clinical terminology and multiple instructions across departments, generating confusion and anxiety. Some patients reported being unable to read written information; others could not access digital systems (e.g., Trust specific patient information apps) due to limited digital skills, lack of devices or difficulty navigating passwords. Unstable housing also meant that letters sent to registered addresses did not reliably reach recipients. Growing reliance on apps, text messaging and online opt-in processes creates additional exclusion for patients with limited technological access or cognitive capacity to manage them. |
| **Transport, logistical and cost barriers to attendance** | Attending multiple diagnostic appointments, often across different hospital sites (for example, CT at one hospital, PET or EBUS at another, treatment at a third), was described as expensive and logistically difficult. Patients reported having to book transport days in advance and waiting hours to be collected, resulting in very long days. For patients already managing mental health conditions, physical fragility and financial precarity, the practical demands of attending at multiple sites functioned as a significant barrier to pathway completion. |
| **Capacity constraints and loss of relational support** | Bottlenecks in key diagnostic procedures (particularly PET scans, biopsies and clinic slots) were identified as sources of delay affecting all patients, but with disproportionate impact on adults with SMI given the motivational, logistical and relational factors affecting their capacity to sustain engagement across multiple touchpoints. Practices that previously supported this group, such as a named clinician accompanying patients to specialist clinics, had been discontinued due to staffing pressures, further reducing relational continuity. |
| **Inconsistent inquiry about mental health in physical health settings** | While routine clinical history covers physical health in detail, mental health history (including current diagnoses, medications and support arrangements) is not consistently recorded or discussed in cancer pathway settings. Staff described patients with past or current mental health conditions going unidentified unless they were on medication or volunteered the information. Without this context, clinicians were unable to anticipate likely barriers, make reasonable adjustments, or involve care coordinators and community mental health teams at the right points in the pathway reducing opportunities to keep patients engaged in the pathway. |
| **System-level barriers** |  |
| **Poor coordination between mental and physical health services** | Staff in physical health settings reported being unable to communicate directly with community mental health teams, relying on indirect routes that were slow and dependent on personal networks. Adults with SMI are more likely than the general population to be managed across multiple services simultaneously (primary care, community mental health, specialist physical health, and sometimes social care or forensic services) and are therefore more likely to encounter gaps at the boundaries between them. In emergency settings, physical symptoms that would otherwise prompt a cancer referral could be deprioritised in favour of immediate mental health management, with no clear mechanism for ensuring follow-up elsewhere. |
| **Unclear accountability for care coordination and re-engagement** | Following missed appointments or failed communications, staff and patients described uncertainty about whether primary care, the mental health team or the specialist cancer service held responsibility for re-initiating contact. This fragmentation was experienced by patients as being “passed around” between services, and by staff as a structural absence of defined accountability. Where engagement was sustained, it was often through the personal effort of specific staff members acting outside formal structures rather than through defined cross-service protocols. Clinicians in physical health settings sometimes assumed that GP follow-up would occur, despite evidence that this population engages poorly with primary care. This was identified as a mechanism contributing to late-stage diagnosis and reliance on emergency presentation routes. |
| **Diagnostic overshadowing** | Staff described a recurring pattern in which physical symptoms in adults with SMI were deprioritised or attributed to psychiatric causes, delaying investigation and referral. Examples included persistent cough being interpreted as another exacerbation of COPD in long-term smokers, and weight loss or fatigue being attributed to medication or mental health conditions rather than triggering further investigation. |
| **Confusing service commissioning and inflexible system design** | Patients described confusion about the nature and legitimacy of services they were referred to, including uncertainty about whether services were part of the NHS or separate organisations operating from NHS premises. This reinforced mistrust at the individual level and acted as a barrier to sustained engagement. Participants with lived experience also identified the NHS more broadly as lacking flexibility and inclusive design, noting that “one-size-fits-all” processes exclude those with complex needs such as difficulties opening mail or answering phones. |
| SMI = severe mental health condition; NHS = National Health Service; GP = General Practice; DNA = Did Not Attend; FDS = Faster Diagnosis Standard; CT = computed tomography; COPD = Chronic Obstructive Pulmonary Disease; PET = positron emission tomography; EBUS = endobronchial ultrasound. |  |

**Table 3.** Enablers and protective factors across the lung cancer screening and diagnostic pathway for adults with severe mental health condition.

| Enablers and protective factors |  |
| --- | --- |
| **Trusting relationships and personally meaningful motivators (individual level)** | Trusting relationships with specific clinicians, support workers, coordinators or peer supporters helped individuals feel reassured and able to navigate complex processes. Relational and personally meaningful motivators (concern for a partner, family member or pet) were identified as important triggers for engagement and behaviour change. When identified and worked with, these individually meaningful motivators were described as more effective than generalised health messaging. |
| **Continuity of care with a named clinician or coordinator (service level)** | Continuity with a familiar clinician across the pathway was repeatedly described as reassuring and stabilising. Examples included nurses accompanying patients to specialist clinics in another hospital so there would be a known face, and single points of contact reducing confusion and anxiety. Continuity was described as particularly important at |
|  | transitions between investigation, diagnosis and treatment, where disengagement risk is highest. |
| **Support workers and patient navigators (service level)** | Mental health support workers, community psychiatric nurses and peer support workers were identified by both staff and patients as central enablers of pathway completion. They helped patients attend appointments, understand correspondence, provided reassurance and maintained contact during periods of instability. Their availability was described as determining whether some individuals completed the pathway at all, though provision was inconsistent across teams. |
| **Adaptive and flexible clinical practice (service level)** | Staff described a range of adaptive practices that supported engagement, including booking follow-up appointments as a failsafe, providing day-before reminders, recording reasonable adjustments on electronic records, using opportunistic moments (bloods, weight, X-ray) to check multiple indicators at once, and persevering with patients who repeatedly missed appointments. These practices were effective but relied heavily on the initiative and experience of particular staff members as they were not embedded in routine service delivery. |
| **“One-stop shop” consolidated diagnostic models (system level)** | Consolidating multiple investigations into a single visit was identified as a promising model by both staff and patients. The approach reduces navigational burden, increases opportunistic case-finding (for example, weight loss noticed during a routine blood test), and addresses the fragmentation of current pathways. Existing examples from breast cancer services were mentioned as potentially adaptable to lung cancer pathways for adults with SMI. |
| **Proactive outreach and peer support (system level)** | Outreach-based approaches that do not rely on patients navigating themselves into services were identified as important for reaching adults with SMI. Embedding lung cancer screening questions within the SMI Annual Physical Health Check was suggested as one route. Peer support workers, seen by patients as relatable and as having been through the process themselves, were identified as enablers of both initial engagement and sustained participation. |
| **Trauma-informed approaches and environmental adjustments (system level)** | Trauma-informed care was identified as important given that many adults with SMI have had traumatic experiences of healthcare services, including previous detentions under the Mental Health Act. Environmental and procedural adjustments (private waiting spaces, reduced sensory stimulation, longer appointment slots, home visits where possible) were described as reducing the likelihood of disengagement and supporting sustained participation. |
| SMI = severe mental health conditions. |  |

**Table 4.** Integrating and consolidating quantitative and qualitative findings using CFIR 2.0.

| # | Consolidated Finding | Quantitative Evidence | Qualitative Evidence | CFIR Domain(s) |
| --- | --- | --- | --- | --- |
| 1 | Adults with SMI represent a small but clinically high-risk population, with higher smoking prevalence, higher lung cancer prevalence, and higher overall cancer rates. | Higher smoking rates; higher lung cancer prevalence; higher total cancer burden; ~3% SMI population living with cancer | — | **Outer Setting**<br>(Population Need & Inequity Context) |
| 2 | Symptom recognition, help seeking, and engagement with screening and diagnostic pathways are reduced in adults with SMI due to interacting individual-level factors (limited awareness and normalisation of symptoms, smoking related stigma and self-blame, fear of diagnosis, avoidance behaviours, and deprivation related hopelessness) | Lower detection rate for lung cancer and elevated DNA rates | Symptom normalisation; minimisation; confusion; reduced help-seeking; fear of diagnosis; stigma; avoidance; deprivation related hopelessness | **Individuals** |
| 3 | The lung cancer pathway is complex, fragmented and insufficiently adapted to the needs of adults with SMI. Cognitive, emotional and practical challenges interact with rigid service processes, lack of reasonable adjustments, and insufficient coordination to increase patient burden and limit pathway navigation and engagement. | Higher DNA rates (lung + all cancers); lower 28-day FDS performance | Fragmented services; difficulty navigating multiple providers; overwhelm; insufficient support; lack of adapted communication; rigid process | **Inner Setting**<br>(Structural Characteristics) + **Innovation** |
| 4 | Higher referral rates do not translate into better outcomes, as adults with SMI are less likely to have lung cancer detected via GP USC referral and less likely to receive a diagnosis within 28 days | Higher suspected referral rates; lower GP-detected lung cancer; lower 28-day FDS performance | Consistent with fragmented services, diagnostic overshadowing, and rigid DNA policies | **Implementation Process** |
| 5 | Higher non-attendance and rigid re-referral processes disproportionately disrupt pathway progression for adults with SMI. | Elevated DNA rates (lung + all cancers) | Rigid non-attendance policies; re-referral barriers | **Inner Setting** |
| 6 | Early detection opportunities are frequently missed due to diagnostic overshadowing and limited proactive investigation during routine contacts. | Lower GP-detected route proportion | Diagnostic overshadowing; missed early investigation | **Implementation Process + Individuals** |
| 7 | A substantial proportion of adults with SMI fall outside NHS Lung Cancer Screening (LCS) programme age eligibility, particularly working age adults with higher smoking prevalence, creating structural gaps in early detection. | LCS programmes have a crucial role in identifying and screening eligible patients. ~One in four adults with SMI in SEL were recorded as current smokers in primary care, although likely underreported. | Consistent with younger age profile discussed by staff; gap between risk accumulation and screening availability | **Innovation + Outer Setting** |
| 8 | Local data systems do not consistently include SMI flags in | Lack of SMI flag; inability to identify | Consistent with staff lacking visibility of SMI | **Inner Setting + Implementation Process** |
|  | cancer reporting, limiting visibility of risk and outcomes. | lung cancer within SMI cancer cohort | status within cancer systems |  |
| CFIR = Consolidated Framework for Implementation Research (CFIR 2.0; Damschroder et al., 2022); SMI = severe mental health condition; NHS = National Health Service; GP = General Practice; DNA = Did Not Attend; USC = Urgent Suspected Cancer; FDS = Faster Diagnosis Standard. |  |  |  |  |

### Individual-level barriers

> *People can get very frightened and worried about going to the GP or about opening up. They are concerned about being judged, about being criticised, about being patronised.* (Participant 1, person with lived experience of SMI)

Four interconnected individual-level barriers were identified and outlined in Table 2. These operated largely before clinical contact and reflected the effects of stigma, fear, limited health literacy, and the weight of competing daily demands on patients already managing mental health condition and social disadvantage. Barriers at this level commonly resulted in symptom concealment, delayed help-seeking, or active disengagement from diagnostic pathways once offered.

### Service-level barriers

> *I was just thinking of an example of a patient who DNA’d… a CT [computed tomography] or PET [positron emission tomography] scan or something… a few times — he was just never going to get there for an early morning appointment. I can remember the consultant actually contacted them and explained the difficulties that this man was having in attending and trying to keep the referral open, because what they do is if somebody DNAs three times, they just close the referral. So, then you have to go back, and you have to re-refer them and they start again.* (Participant 2, clinical staff member)

Five service-level barriers were identified (Table 2). These reflected a pathway whose structure, communication practices and logistical demands were poorly adapted to the needs of this group, with rigid administrative process, complex correspondence and fragmented access arrangements. Engagement frequently faltered at multiple points in the pathway. Barriers at this level typically affected adults after referral, contributing to delayed progression through the pathway.

### System-level barriers

> *Everything has to go via PCMHT [primary care mental health teams] who are like, “we don’t know what’s going on”, and you’re like, well, that’s super helpful. But I need to speak with that person over there because I’m seeing them and I’m worried. And so, it’s more like whether you’re lucky enough to know what their coordinator’s name is and their phone number, and then you have to go out of your way to contact them.* (Participant 3, clinical member of staff)

Four system-level barriers were identified (Table 2). These reflected structural features of how physical and mental health services are organised and the limited coordination between them across SEL. Poor coordination between services, unclear accountability for care continuity, and clinical assumptions deprioritised physical symptoms in adults with SMI. Where engagement across services did occur, it was often through the personal effort of specific staff instead of through embedded cross-service protocols.

### Enablers and protective factors

> *Because they don’t know the hospital. They don’t know any faces there. Then they see you and think, “oh good, it’s [name]”. They feel a bit more reassured… and it’s really sad that we’re not doing that at the minute because I think it really helped people.* (Participant 4, staff member)

Although barriers predominated, seven enabling factors were identified across all three levels (Table 3). Trusting relationships, continuity with named clinicians and support workers, and adaptive clinical practice emerged as the most consistently described facilitators of engagement, alongside system-level approaches such as consolidated diagnostic models and proactive outreach. These enablers were largely informal and inconsistently available, as they relied mainly on individual initiative and not on routine service delivery.

### Integration of findings

Integration of quantitative and qualitative data produced eight consolidated findings mapped on CFIR that together characterise a consistent pattern of pathway inequality. The quantitative data established that adults with SMI have higher baseline lung cancer risk, higher referral rates, but poorer post-referral outcomes including lower detection rates, poor FDS compliance, with longer time to diagnosis and higher non-attendance across the pathway. The qualitative data explained that stigma and mistrust suppress symptom disclosure before referral; fear and avoidance reduce engagement with investigations once referred; rigid service processes disproportionately disadvantage people whose attendance is inherently less predictable due to social and clinical realities of living with SMI; and fragmented system structures fail to provide the coordination needed to sustain engagement across a complex, multi-step pathway. Crucially, the integration revealed that the inequality does not arise at a single point of failure (e.g. at screening) but accumulates across sequential pathway stages, compounding at each transition point along the pathway.

## Discussion

This study provides evidence of inequalities across the lung cancer pathway for adults with SMI in SEL. The findings extend the existing evidence base which has largely focused on demonstrating excess cancer mortality in SMI populations or reduced screening uptake (Chen et al., 2024, Cancer Research UK, 2025, Tredget and Williams, 2023), by characterising where along the diagnostic pathway these inequalities arise and identifying the mechanisms through which they operate. This granularity is important for informing targeted, proportionate interventions for this vulnerable population.

A central finding of this study is that inequalities in lung cancer outcomes for people with SMI are not due to poor access to referral opportunities, but instead limited pathway adherence and diagnostic delays that perpetuate poorer outcomes. For example, the USC referral rates were substantially higher in the SMI population, consistent with higher primary care consultation rates and clinical complexity (Kontopantelis et al., 2015). However, high referrals did not result in higher rates of diagnosis for referred patients, highlighting more may be needed beyond the point of referral to support SMI patients once referred to cancer services. Positively, a range of programmes are already in place locally, including annual physical health checks, tobacco dependency and cessation programmes, and the former Targeted Lung Health Checks pilot, which has recently been accepted to become a national LCS Programme, which provide an opportunity to consider better approaches to integrating care along the diagnostic pathway for the SMI population.

Another key finding was that adults with SMI were less likely to have lung cancer detected via GP referral (which is the most optimal route for continuity of care) despite higher overall referral rates, and no SMI lung cancer diagnoses met the 28-day FDS, indicating greater reliance on emergency presentation. This points to concentrated disadvantage after referral. Patients may disengage or be lost between services through missed appointments, while diagnostic overshadowing can delay investigation when physical symptoms are attributed to psychiatric causes, medication, smoking or clinical complexity, or are concealed by patients because of stigma. Improving referral and diagnostic rates alone is therefore unlikely to address these mechanisms. Equitable progress requires attention to routes to diagnosis, pathway navigation, engagement and coordination across providers, alongside training and structural changes. For example, embedding routine physical-health checks in mental health settings and alerting cancer pathway staff to patient’s mental health needs at the point of referral (Shefer et al., 2014, Health Services Safety Investigations Body, 2025).

The qualitative findings further illustrate these patterns. Individual-level barriers such as stigma, fear, low health literacy, competing survival priorities, help explain why adults with SMI may be less likely to present symptoms or sustain engagement once referred (Clement et al., 2015, Carter-Harris, 2015, Grassi and Riba, 2021).

Service-level barriers, such as rigid DNA policies, inaccessible correspondence and inconsistent support worker availability illustrate how pathway processes that function adequately for the general population can create significant obstacles for people with complex needs (Mitchell and Selmes, 2007). System-level barriers such as fragmented physical and mental health care coordination, diagnostic overshadowing and unclear care coordination responsibility reveal structural features of healthcare organisation that undermine equitable access despite individual staff efforts (Thornicroft et al., 2022, Shefer et al., 2014). Inequality for adults with SMI does not arise at a single point of failure. Instead, it accumulates across multiple post-referral pathway stages. Among those who do develop lung cancer, fewer are diagnosed via GP referral for suspected cancer routes and more rely on alternative or unmanaged pathways. Post-referral, diagnostic timeliness and engagement are noticeably poorer (Cunningham et al., 2024, Virgilsen et al., 2022).

The screening eligibility findings raise a distinct but related challenge. The NHS LCS programme appropriately targets people aged 55–74 who have ever smoked, and the data confirms that two-thirds of lung cancers in the SMI population do occur within this eligible age group. However, a high number of people with SMI in SEL currently do not meet the eligibility criteria for LCS, despite higher smoking rates, substantial tobacco exposure and, more specifically, living in deprived areas place people with SMI at greater risk of developing lung cancer (Bernat et al., 2012, Chen et al., 2024). This creates a gap between when risk begins to accumulate in this population and when screening-based detection becomes available and suggests the LCS programme is a critical opportunity to detect lung cancer earlier if SMI patients can be supported to effectively participate in screening. Complementary prevention and earlier risk identification strategies could help bridge this gap to identify and support younger SMI patients at rising risk of developing lung cancers without requiring changes to the national screening criteria. Although smoking prevalence was approximately six-fold higher among adults with SMI, this study could not directly examine the interrelationship between smoking exposure, lung cancer and SMI, as pack year and initiation data were not available. This triangulation, however, warrants dedicated investigation. Smoking cessation is a modifiable, potentially high-impact lever, and embedding cessation support within existing touchpoints such as the APHC could help reduce future lung cancer burden, particularly in the under 55 cohort who currently fall outside NHS LCS screening eligibility. Additionally, increasing national reporting of cancer outcomes for adults with SMI is needed if local systems are to understand and address disparities in lung cancer for patients with SMI whilst implementing the LCS programme.

Finally, CWT metrics are not routinely stratified by SMI status, meaning that the inequalities documented here would not be visible in standard performance reporting. Without measurement, these disparities risk remaining unmanaged. The introduction of an SMI flag within cancer tracking systems across primary and secondary care would represent a relatively low-cost intervention with significant potential to improve visibility and accountability of mitigating rising risk in the SMI population, consistent with the Core20PLUS5 principles (NHS England, 2021).

### Strengths and limitations

This study integrates quantitative pathway data with qualitative insight from both staff and people with lived experience, providing a more comprehensive understanding of lung cancer inequalities for adults with SMI than either approach could achieve alone. The multi-stakeholder validation process, which involved clinicians, people with lived experience, and system partners, strengthened the credibility and implementation relevance of the findings. The study addresses a recognised evidence gap in understanding how national cancer pathway standards operate in practice for adults with SMI at a local level.

Several limitations should be noted. Firstly, the quantitative dataset used for this study, focuses on patients recorded on SMI registers within GPs in SEL in a specific time period, it is accepted that this may not represent the totality of the SMI population in SEL, only those that are registered with their GP as having an SMI diagnosis and so should be interpreted as such. The recorded smoking prevalence (27%) sits well below estimates from inpatient and national samples (commonly ≥ 60%), likely reflecting under-recording in primary care QOF registers. How smoking status is captured and updated in primary care for this population warrants further investigation, particularly as NHS LCS programme eligibility draws on primary care smoking records. Such under-recording may leave eligible adults with SMI unidentified and uninvited. Self-reported surveys and national reports typically yield higher estimates. As pack-year history and smoking initiation age were not available in the dataset, it is difficult to quantify directly the cumulative tobacco exposure in working-age SMI adults. The quantitative data are cross-sectional and descriptive, rates are not age-standardised, and small numbers in the SMI cohort (particularly for lung cancer-specific outcomes) limit precision and preclude robust statistical testing, for example as depicted in Figure 2, limited data availability on lung cancer prevalence rates for SMI patients in Bromley and Lambeth resulted in high variability reporting across SEL boroughs. The available dataset did not include formal Routes to Diagnosis coding, tumour stage at diagnosis, pre-referral intervals, or survival outcomes, restricting the ability to quantify the contribution of specific pathway intervals to observed delays. Qualitative data are drawn from a single geographical area and may not be directly generalisable to other settings, though many of the barriers identified reflect widely recognised features of the NHS landscape. The sequential study design does not permit causal inference, and findings are interpretative and explorative. Qualitative participants were not sampled based on their quantitative pathway data, and direct linkage between individual patient journeys and pathway metrics was not possible.

## Conclusions

Adults with SMI in SEL experience a consistent and cumulative pattern of inequality across the lung cancer pathway, from higher baseline risk through to poorer diagnostic pathway performance. The evidence demonstrates that the inequality does not arise at a single point of failure but accumulated across sequential pathway stages. A substantial number of adults with SMI fall outside screening eligibility and despite lung cancer prevalence being high in the 55-74 SMI cohort, diagnosis rates remain low for this population. Fewer cancers are detected via GP referral for suspected cancer routes and post-referral, diagnostic timeliness, and engagement are noticeably poorer. Addressing these inequalities will require coordinated action across prevention, identification, pathway navigation, diagnostic timeliness, and routine measurements. By generating practical, system-level evidence on where lung cancer pathways fail for adults with SMI, this study provides a foundation for moving from awareness of inequality to structured, accountable actions that address barriers to accessing and adhering to lung cancer pathways for the SMI population.

## Data Availability Statement

The raw data supporting the conclusions of this article will be made available by the authors, without undue reservation.

## Ethics statement

This study received Health Research Authority (HRA) from the Research Ethics Committee (REC) under 25/LO/0523 in 2025. Additional approvals for this study were obtained from the Clinical and Effectiveness Governance Committee and Information Governance Committee at South London and Maudsley NHS Foundation Trust prior to implementation. All participants provided their written informed consent to participate in this study.

## Author contributions

GT and JWi: designed the mixed-methods study. JWi and WP: obtained the HRA approval for the study. GT: led the study on behalf of the project team. GT: led the quantitative data collection, analysis and write up of quantitative results with support from DI’A and JWi and developed the integrated findings framework. RP: led the qualitative data collection (i.e., interviews and focus groups) and workshops to co-define and validate the findings with system stakeholders, with support from GT, MM, WP, DK and CC. MM: led the qualitative analysis and write up of qualitative results with support from RP, WP, GT and SS. The manuscript was written by MM and GT, and refined by JWi, RP, DI’A, KH, WP, SG and ME. All listed co-authors have contributed to the article and approved the final version submitted for publication.

## Acknowledgements

The authors would like to thank all participants who took part in the evaluation, as well as the people with lived experience and clinical staff who supported its completion. The authors wish to extend thanks to all colleagues that participated in the Steering Group for this project, including our core partners from South London & Maudsley NHS Foundation Trust, Guy’s & St Thomas’ NHS Foundation Trust, King’s College Hospital and King’s College London. Particular thanks to our Clinical Care and Professional Leadership representative for SEL, Mark Adams, and David I’Anson and the SEL ICB Business Intelligence team, which in partnership with the ICB Care Group Leads for planned care and cancer, have produced invaluable dashboards and insights which have made this work possible.

## Supplementary materials

Please see separate file included with submission.

