## Supplementary files 1-4 for "Lung cancer pathway inequalities for adults with severe mental health conditions: A mixed-methods analysis of barriers to screening and care pathways in South East London"

**Supplementary File 1: Project Steering Group stakeholder participation list by type of organisation and type of role**

| **Organisational type** | **Role type** | **No. representatives** | |
| --- | --- | --- | --- |
| Clinical academic partner | Leadership | 5 | |
|  | Delivery team | 3 | |
|  | Research | 2 | |
| Academic partner | Clinical and academic cancer specialists | 2 | |
| Primary care partner | General practitioners | 3 | |
|  | Clinical/frontline staff (nurse) | 1 | |
| Secondary care partner (mental health) | Clinical/frontline staff | 8 | |
|  | Operational management | 3 | |
|  | Senior director/leader | 4 | |
| Acute care partner | Clinical/frontline staff | 8 | |
|  | Operational management | 1 | |
|  | Senior director/leader | 1 | |
| Voluntary, Community and Social Enterprise Partner | Senior director/leader | 3 | |
|  | Operational management | 1 | |
| Lived experience expertise | Expert by experience of severe mental health conditions | 2 | |
| Project funder and co-lead | Programme management | 9 | |
| Mental health commissioner / Integrated Care Board | Operational management | 4 | |
| Professional body | Senior director/leader | 1 | |
| Local authority / public health partner | Operational management | 1 | |
| **Total participants** | | | 62 |

Wider stakeholder groups have included:

- King’s Health Partners Cancer & Respiratory Clinical Academic Groups
- NHS England – London Physical Health & SMI Community of Practice
- SEL Lung Cancer & SMI Community of Practice
- Pan London Physical Health Leads Network
- Cancer Research UK Research Advisory Group for Lung Cancer Screening for People with SMI

**Supplementary file 2: Integrated coding framework outlining themes and sub-themes.**

| Dataset | Domain (CFIR) | Sub-theme | Sub-theme | TDF Domain |
| --- | --- | --- | --- | --- |
| Interviews with Staff | Innovation | Innovation design; Innovation complexity | “One-stop shop” consolidated diagnostic models | Environmental context & resources; Behavioural regulation |
|  | Outer Setting | Partnership & connections | Fragmented care systems, lack of integration (with mental health services) | Social influences; Environmental context & resources |
|  |  |  | Unclear accountability for care coordination and re-engagement | Social/Professional roles & identity; Environmental context & resources |
|  |  |  | Referral and communication pathways | Environmental context & resources |
|  |  | Local conditions | Financial, logistical and transport barriers to access | Environmental context & resources; Beliefs about consequences |
|  |  |  | Local population context and variation in SMI presentation | Knowledge; Beliefs about consequences; Environmental context & resources |
|  |  | Local conditions; Policies & laws | Confusing service commissioning and inflexible system design | Environmental context & Resources; Beliefs about consequences |
|  | Inner Setting | Structural characteristics: work infrastructure | Rigid non-attendance (Did Not Attend – DNA) and re-referral policies | Environmental context & resources; Behavioural regulation |
|  |  |  | Staffing and responsibilities | Social/Professional roles & identity; Social influences |
|  |  | Structural characteristics: work infrastructure; available resources | Staffing, workload, high staff turnover | Environmental context & resources; Social/Professional roles & identity |
|  |  | Available resources; Relational connections | Capacity constraints and loss of relational support | Environmental context & resources; Social/Professional roles & identity |
|  |  | Relational Connections; Structural characteristics; work infrastructure | Rigid service design and lack of interdisciplinary teams | Social/Professional roles & identity; Skills |
|  |  | Culture: Recipient-centredness; Culture: human equity-centredness | Diagnostic overshadowing | Knowledge; Skills; Beliefs about consequences; Social/Professional roles & identity |
|  |  | Relational connections | Staff attitudes & relational continuity (familiarity with a names clinician) | Social/Professional roles & identity; Social influences; Beliefs about consequences |
|  |  | Culture: Recipient-centredness; Structural characteristics: physical infrastructure | Trauma-informed approaches and environmental adjustments | Skills; Environmental context & resources; Emotion; Beliefs about consequences |
|  | Individuals | Innovation recipients: capability; motivation | How people with SMI present to services | Beliefs about capabilities; Behavioural regulation; Social influences |
|  |  | Innovation recipients: need | Co-occurring substance use and risk profile | Social influences; Beliefs about consequences |
|  |  | Innovation recipients: opportunity; motivation | Readiness of competing priorities at point of engagement | Environmental context & resources; Emotion; Memory, attention & decision processes |
|  |  | Innovation recipients: need: capability | Interplay between physical and mental health / impact of mental health conditions | Emotion; Beliefs about consequences |
|  |  | Innovation recipients: capability; opportunity | Digital exclusion | Environmental context & resources |
|  | Implementation Process | Engaging: Innovation recipients | Role of support worker / patient navigator | Social/Professional roles & identity; Social influences; Skills; Environmental context & resources |
|  |  |  | Continuity of engagement across care phases / follow up | Social/Professional roles & identity; Behavioural regulation; Beliefs about consequences; Environmental context & resources |
|  |  |  | Late presentation and missed detection pathways | Beliefs about consequences; Environmental context & resources; Social influences |
|  |  |  | Proactive outreach and peer support | Social influences; Environmental context & resources; Reinforcement |
|  |  | Tailoring strategies; Adapting | Adaptive engagement and flexible clinical practice | Skills; Behavioural regulation; Social/Professional roles & identity |
|  |  | Assessing context | Clinical prioritisation (physical symptoms being deprioritised) | Knowledge; Skills; Beliefs about capabilities |
|  |  | Assessing needs: Innovation recipients | Inconsistent routine inquiry and disclosure of mental health history | Skills; Memory, attention & decision processes; Social influences |
|  | Equity & Social Determinants (HEIF overlay) | HEIF overlay with Innovation > Innovation recipients: Motivation; Outer Setting > Local attitudes | Stigma / comparative perceptions | Social influences; Emotion: Social/Professional roles & identity; Beliefs about consequences |
|  | Equity & Social Determinants (HEIF overlay) | HEIF overlay with Outer Setting > Local conditions; Individuals > Innovation recipients: Opportunity/Need | Social vulnerability and living environment | Environmental context & resources; Social influences |
|  | Equity & Social Determinants (HEIF overlay) | HEIF overlay with Inner Setting > Culture: Recipient-Centredness; Individuals > Innovation recipients: Motivation | Mistrust / perceived coercion in healthcare interactions | Beliefs about consequences; Social/Professional roles & identity; Emotion |
| Focus groups with adults with lived experience of SMI | Innovation | Innovation design | Coercive, intrusive or fear-based messaging makes people hide | Social influences; Behavioural regulation; Emotion |
|  | Outer Setting | Partnerships & connections | Access and referral pathways / delayed escalation | Environmental context & resources |
|  |  |  | Unclear referral routes | Knowledge |
|  |  | Local conditions | Inflexible and inaccessible systems | Environmental context & resources |
|  |  | Local conditions; Policies & laws | Confusing service commissioning | Environmental context & resources; Beliefs about consequences |
|  |  | Local attitudes | Cultural attitudes | Environmental context & resources; Social influences |
|  | Individuals | Innovation recipients: Motivation | Ambivalence / enjoyment of smoking | Reinforcement; Emotion; Beliefs about consequences |
|  |  |  | Motivation triggers (personally meaningful motivations) | Goals; Reinforcement; emotion; Social influences |
|  |  | Innovation recipients: capability; needs | Interplay between physical and mental health / impact of mental health conditions | Emotion; Beliefs about consequences |
|  |  | Innovation recipients: capability; opportunity | Digital exclusion | Environmental context & resources |
|  | Implementation process | Engaging: Innovation recipients | Need for advocacy / hand holding (need for navigation support ) | Behavioural regulation; Social influences; Social/Professional roles & identity |
|  |  |  | Communication and delivery of care | Skills; Knowledge; Emotion; Beliefs about consequences |
|  |  |  | Proactive outreach / peer support | Social influences; Environmental context & resources; Reinforcement |
|  | Equity & Social Determinants (HEIF overlay) | HEIF overlay with Individuals > Innovation recipients: Motivation; Outer Setting > Local attitudes | Feeling judged / fear /stigma | Social influences; Beliefs about consequences; Emotion |
|  |  | HEIF overlay with Individuals: Innovation recipients: capability; Inner setting > communications | Health literacy and gaps/barriers in communication | Knowledge; Skills |
|  |  | HEIF overlay with Inner Setting > Culture: Recipient-Centredness; Individuals > Innovation recipients: Motivation | Trust and perceived responsiveness of primary and secondary care services | Beliefs about consequences; Emotion |
| CFIR = Consolidated Framework for Implementation Research (Damschroder et al., 2022); HEIF = Health Equity Implementation Framework (Woodward et al., 2021); TDF = Theoretical Domains Framework (Cane et al., 2012); SMI = severe mental health conditions (also referred to as severe mental illness); | | | | |

**Supplementary file 3: Distribution of adults with severe mental health condition (SMI) by age band, South East London**

| **Age band** | **Non-SMI population (n)** | **SMI population (n)** | **Total population (n)** | **% of age band with SMI** |
| --- | --- | --- | --- | --- |
| 0–19 | 439,476 | 214 | 439,690 | 0.05% |
| 20–39 | 703,508 | 7,558 | 711,066 | 1.06% |
| 40–54 | 399,417 | 8,878 | 408,295 | 2.17% |
| 55–74 | 329,126 | 6,516 | 335,642 | 1.94% |
| >74 | 147,005 | 3,199 | 150,204 | 2.13% |
| **All ages** | **2,018,532** | **26,365** | **2,044,897** | **1.29% overall SMI prevalence** |

Counts represent patients recorded on practice SMI registers (Quality and Outcomes Framework definition). Percent SMI calculated as SMI population divided by total population within each age band. South East London aggregate.

**Supplementary file 4: Participant characteristics**

| **Clinical healthcare staff (interviews)** | | |
| --- | --- | --- |
| **Service Area** | **Role type** | **No. of participants** |
| Primary Care | General Practitioner | 2 |
| Emergency Department | Emergency Department Consultant | 1 |
| Secondary Care | Lung Clinical Nurse Specialist | 6 |
|  | Smoke Free Services | 1 |
|  | Mental Health Nurse | 1 |
| **Adults with lived experience of severe mental health conditions (focus groups)** | | 6 |
| **Total number of participants** | | 17 |
